# Pilot study of the FoxP3+ T lymphocyte count and its relationship with the Gensini score in patients with coronary heart disease and type 2 diabetes mellitus from a South American cohort

**DOI:** 10.1101/2023.10.08.23296720

**Authors:** María Juliana Estévez Gómez, Diego Andrey Acevedo Peña, Silvia Fernanda Castillo Goyeneche, Anderson Felipe Arias, Luis Andres Dulcey Sarmiento, Jaime Gomez, Carlos Hernandez, Juan Sebastián Theran Leon, Raimondo Caltagirone, Edgar Blanco, María Ciliberti

## Abstract

**INTRODUCTION:** One of the pathogenic links between type 2 diabetes mellitus (T2DM) and coronary heart disease (CHD) is low-intensity chronic inflammation, which is limited by FoxP3+ regulatory T cells.

**MATERIALS AND METHODS:** A comparative, observational, single-center, single-stage study was carried out. The severity of atherosclerosis was assessed by calculating the Gensini score based on selective coronary angiography. The absolute and relative content of CD4+CD25 high FoxP3+ and CD4+CD25 low FoxP3+ T cells in the blood was evaluated by flow cytometry.

**RESULTS:** 57 patients with chronic ischemic heart disease were examined. Patients with a combination of CHF and T2DM are characterized by an increase in the relative and absolute content of FoxP3+CD25 lymphocytes in the peripheral blood and an increase in the intranuclear content of FoxP3 in them, which is more pronounced in patients with disease moderate. atherosclerosis (Gensini score 17 to 45 points).

**CONCLUSIONS:** This was the first Latin American study that managed to show the relationship between an increase in the content of low FoxP3+CD25 lymphocytes in the peripheral blood with atherosclerosis in patients with a combination of coronary artery disease and DM2.

## INTRODUCTION

Atherosclerosis is a chronic disease characterized by lipid accumulation and inflammation in the vessel wall. The development of atherosclerosis and diabetes mellitus (DM) is interconnected at the cellular, molecular and epigenetic levels. At the same time, patients with diabetes are characterized by a complicated and rapidly progressive course of atherosclerosis [1]. Patients with poor glycemic control are at higher risk of developing vascular events than those who achieve target glycemic levels [2].

Low-intensity chronic inflammation is known to be a common pathophysiological feature of both atherosclerosis and type 2 diabetes (T2DM) [1]. Considering that a number of therapeutic approaches aimed at suppressing inflammation have been shown to be effective in reducing cardiovascular risk in patients with and without diabetes [3], it is important to understand the development patterns of inflammation in atherosclerosis and their relationship with the metabolic status of the patients.

Of interest are regulatory T lymphocytes (Treg) that, due to various intercellular interactions, secretory activity and metabolic reprogramming, can limit inflammation and the development of atherosclerosis [4]. As atherosclerosis progresses, Treg cells accumulate in the plaque, where they regulate complex intercellular relationships, including suppressing the activity of proinflammatory T cell subsets, changing macrophage phenotype, reducing the efficiency of antigen presentation, stimulation of smooth muscle cell proliferation and increased collagen production [5].

According to meta-analysis data, patients with T2DM were characterized by a decrease in the content of CD4+CD25+Foxp3+ Treg lymphocytes, which was further aggravated in the presence of macro- and microvascular complications [6]. However, in the analyzed works only the relative content of Treg lymphocytes is studied, without calculating their absolute number of both regulatory and non-regulatory lymphocytes.

FoxP3 is the main transcription factor of Tregs and is necessary for their maturation and immunosuppressive function. Their localization within the cell has been shown to vary and nuclear translocation is an integral attribute of normal Treg activity [7]. Currently, there is no information in Latin America on the effect of metabolic disorders on the level of FoxP3 transfer to the cell nucleus in patients with a combination of atherosclerosis and DM2.

Recently, more and more information has appeared about the population of FoxP3+ T lymphocytes, which practically do not carry CD25 molecules in their membrane, or are present only in small quantities (FoxP3+CD25 or CD4+ T lymphocytes) [7] [8]. The number of these cells increased in patients with autoimmune diseases such as type 1 diabetes (T1D) and systemic lupus erythematosus [8]. It has been suggested that FoxP3+CD25 T cells may represent a new cellular marker of recent autoimmune or cytokine-mediated inflammatory responses in tissues.

## PURPOSE OF THE STUDY

The objective of this study was to study the content of FoxP3+CD25 high and FoxP3+CD25 low T lymphocytes, the proportion of cells with intranuclear localization of FoxP3 and the production of the main regulatory cytokines in relation to clinical and metabolic parameters in patients with a combination of coronary artery disease (CHD) and T2DM compared to patients with coronary artery disease without diabetes.

## MATERIALS AND METHODS

The study was carried out at the Cardiology Research Institute of the National Medical Research Center of the University Hospital of the Andes in Merida Venezuela during 2015 with the support of Harvard University. To evaluate the properties of FoxP3+ T cells, two patient samples were formed: the main group and the comparison group. The main group was formed taking into account the following criteria.

### Inclusion criteria

- Men and women between 40 and 70 years old;
- Chronic ischemic heart disease with stable class II-III angina pectoris;
- Indications for selective coronary angiography;
- DM2.

### Exclusion Criteria

- Any acute cardiovascular complication or coronary artery bypass surgery suffered less than 6 months ago;
- Any acute inflammatory disease suffered less than 1 month ago;
- Serious concomitant pathology or cancer;
- DM
- Refusal to participate in the study.

The comparison group was formed taking into account the following criteria:

- Men and women between 40 and 70 years old;
- Chronic ischemic heart disease with stable angina pectoris class II-III;
- Indications for selective coronary angiography.
- DM1 and DM2 as a diagnosis excluded entry into the control group.
- Any acute cardiovascular complication or coronary artery bypass surgery suffered less than 6 months ago;
- Any acute inflammatory disease suffered less than 1 month ago;
- Serious concomitant pathology or cancer;
- Refusal to participate in the study.

### Method to form a sample from the population under study

The samples were formed by continuously including observations. The study was an observational, single-center, one-stage comparative study. The diagnosis of T2DM was established according to the criteria of the modern classification of DM [9]. All patients underwent angiography using the angiographic complex Artis one and the Digitron-3NAC computing system (Siemens Shenzhen Magnetic Resonance Ltd., Shenzhen, China). The severity of coronary atherosclerosis was evaluated by calculating the Gensini Score.

Venous blood was drawn in the morning on an empty stomach and 2 hours after a standard meal load. Mononuclear leukocytes were obtained from heparinized blood by density gradient centrifugation (Histopaque 1077, Sigma Aldrich, USA). To evaluate the content of FoxP3+ T cells, cells were stained with fluorochrome- labeled monoclonal antibodies : anti- CD4-FITC, anti-CD25-APC, anti-FoxP3-PE (Becton Dickinson, USA) in accordance with the manufacturer’s recommendations. Samples were analyzed on a FACSCalibur flow cytometer (Becton Dickinson, USA) using CellQuestPro software (BD Biosciences, USA). The number of cells with FoxP3 translocation to the nucleus was evaluated by flow cytometry with visualization (Amnis device FlowSight (Luminex, USA)) with the addition of dye-coupled monoclonal antibodies to the panel. 7-Aminoactinomycin D (7-AAD) for nuclear staining. Results were expressed as a percentage of positively stained cells of the desired cell population. The absolute number of cells in the peripheral blood was calculated according to the general blood test.

Peripheral blood mononuclear leukocytes were incubated in complete RPMI medium (Sigma, USA) at a concentration of 10 6 cells per 1 ml of medium at 37 °C and 5% CO 2. Spontaneous cytokine secretion was evaluated and lipopolysaccharide (LPS) (10 μg /ml; Sigma, USA). The supernatant liquid of the cell cultures was collected after 24 hours and the determination of the content of cytokines and chemokines in the blood serum and the supernatant liquid of the cell cultures was carried out with the equipment of the Collective Use Center “Medical Genomics” of the institute of immunology from the University of the Andes in Merida Venezuela. Multiplex analysis was carried out on a FLEXMAP 3D instrument (Luminex Corporation, USA) using the MILLIPLEX map Human Cytokine / Chemokine Panel I kit for the determination of 38 analytes according to the manufacturer’s recommendations and MILLIPLEX Analyst 5.1 software (MERCK, Millipore, Milliplex ; USA).

The glucose concentration was determined by the hexokinase method. The percentage of glycated hemoglobin (HbA 1c) in blood was determined using the immunoturbidimetric method (DiaSys, Germany). Insulin content in blood serum was determined by enzyme immunoassay (AccuBind, USA). The lipid spectrum of blood was studied (the content of total cholesterol (TC), triglycerides (TG), high-density lipoprotein cholesterol (HDL-C), low-density lipoprotein cholesterol (LDL-C), C -LDL/HDL -C ratio) (Kits from JSC “Diakon -DS”, Russia). The degree of insulin resistance was determined by calculating the HOMA index (Homeostasis Model Assessment; fasting glucose × fasting insulin /22.5) and calculating the TG/glucose ratio (Ln (TG (mg/ dL) × glucose fasting (mg/ dL)/ 2) [10].

### Statistic analysis

Statistical processing of the obtained data was performed using the STATISTICA 10.0 software package (StatSoft, USA). To check the normality of the distribution of quantitative indicators, the Shapiro-Wilk test was used. The results were presented as median and interquartile range (Me [Q1; Q3]). The statistical significance of differences in quantitative indicators in independent groups of patients was evaluated using the Mann-Whitney U test. Frequencies of occurrence in independent groups of patients were compared using Pearson’s χ^2^ test or Fisher’s exact test. To evaluate the relationship between the characteristics, Spearman ‘s rank correlation coefficient (Rs) was used. The differences were considered statistically significant at a significance level of p<0.05.

### Ethical review

The study protocol was approved at a meeting of the Biomedical Ethics Committee of the Cardiology Research Institute of the Hospital Universitario de los Andes in Merida Venezuela.

## RESULTS

The study included 57 patients with documented chronic coronary artery disease, including 22 patients with T2DM and 35 patients without DM, who formed the comparison group. In a first stage, we studied the content of FoxP3+ cells and cytokine production in 37 patients. In the second stage of the study, an analysis of the translocation of the transcription factor FoxP3 to the nucleus was carried out in 34 patients. Patient characteristics are presented in the table. 1 and 2.

All patients were receiving regular cardioactive therapy approaching the optimal level. The blood pressure (BP) level during treatment did not exceed 140/90 mmHg. There were no significant differences between groups in the use of the main groups of cardioactive drugs. Antihyperglycemic therapy in patients with DM2 included biguanides (86.4%), and gliptins (13.6%).

In independent groups of patients studied to determine the content of FoxP3+ cells in the blood, the quantitative proportions between men and women were not comparable: among patients with T2DM there were 3 men and 14 women, and among patients with coronary artery disease Without DM2 there were 12 men and 8 women (Table 1), in addition the women were characterized by having a higher body mass index (BMI). In the patient groups in which we studied nuclear transfer of the FoxP3 factor, patients with T2DM were slightly older than patients without diabetes (Table 2). The observed differences correspond to patterns observed in the population as a whole [10], therefore, in a subsequent analysis, we corrected the FoxP3+ lymphocyte indicator for existing intergroup differences in gender, age and BMI.

**Table 1.** Clinical characteristics of the patient groups for the analysis of FoxP3+ cell content and cytokine production.

| Variables | Patients with a combination of coronary artery disease and T2DM.<br>(n=17) | Patients with coronary artery disease without diabetes.<br>(n=20) | p value |
| --- | --- | --- | --- |
| Gender (male/female), n (%) | 3/14 (17.6/82.4) | 12/8 (60.0/40.0) | 0.018 |
| Year old | 65.0 [62.0; 68.0] | 64.0 [58.0; 66.0] | 0.220 |
| Duration of IC, years | 5.0 [3.0; 10.5] | 4.0 [ 2.0; 8.0] | 0.305 |
| Duration of T2DM, years | 9.5 [3.0; 12.0] | - | - |
| Duration of hypertension, years. | 18.0 [10.5; 35.0] | 15.0 [5.0; 21.0] | 0.182 |
| Systolic blood pressure, mm Hg. Art. | 134.0 [115; 140] | 123.0 [112; 134] | 0.257 |
| Diastolic blood pressure, mm Hg. Art. | 70.0 [64.0; 78.0] | 77.0 [65.0; 80.0] | 0.257 |
| Smoking, n (%) | 2 (11.7) | 7 (35.0) | 0.137 |
| BMI, kg/ m<sup>2</sup> | 31.6 [29.7; 34.6] | 29.4 [26.2; 31.1] | 0.022 |
| Waist circumference, cm | 103.8 [97.5; 117.5] | 98.0 [93.0; 106] | 0.142 |
| Gensini score , points | 26.5 [15.0; 56.5] | 16.8 [2.5; 38.0] | 0.311 |
| Statins , n (%) | 14 (82.4) | 16 (80) | 0.998 |

**Note.** IHD - coronary heart disease; DM - diabetes mellitus; AH - arterial hypertension; BMI: body mass index.

**Table 2.** Clinical characteristics of the patient groups for the analysis of the translocation of the FoxP3 factor to the nucleus.

| Variables | Patients with a combination of coronary artery disease and T2DM.<br>(n=12] | Patients with coronary artery disease without diabetes.<br>(n=22] | Relative risk |
| --- | --- | --- | --- |
| Gender (male/female), n (%) | 7/5 (58.3/41.7) | 18/4 (81.8/18.2) | 0.224 |
| Year old | 67 [61.5; 70] | 61 [55; 66] | 0.049 |
| Duration of IC, years | 8 [3; 9] | 3 [ 2; 9] | 0.309 |
| Duration of T2DM, years | 6 [3; 10.5] | - | - |
| Duration of hypertension, years. | 17.5 [11; twenty] | 16 [15; twenty] | 0.958 |
| Systolic blood pressure, mm Hg. Art. | 134 [108; 149] | 128 [120; 137] | 0.842 |
| Diastolic blood pressure, mm Hg. Art. | 70 [65; 78] | 77 [65; 82] | 0.220 |
| Smoking, n (%) | 3 (25) | 11 (50) | 0.266 |
| BMI, kg/ m<sup>2</sup> | 30.7 [26.5; 33.3] | 28.0 [25.7; 32.4] | 0.423 |
| Waist circumference, cm | 105.0 [96.0; 113] | 100.5 [93.0; 112.0] | 0.749 |
| Gensini score , points | 30.5 [15.0; 46.0] | 30.5 [15.0; 46.0] | 0.254 |
| Statins , n (%) | 10 (83.3) | 19 (86.4) | 0.998 |

**Note.** IHD - coronary heart disease; DM - diabetes mellitus; AH - arterial hypertension; BMI: body mass index.

The group of patients with T2DM was characterized by higher values of the TG/glucose ratio, which indirectly reflects the level of insulin resistance, compared to the group of patients with coronary artery disease without diabetes (Table 3).

**Table 3.** Basic parameters of carbohydrate and lipid metabolism.

| Variables | Patients with a combination of coronary artery disease and T2DM.<br>(n=22) | Patients with coronary artery disease without diabetes.<br>(n=35) | p value |
| --- | --- | --- | --- |
| Fasting glucose, mmol /l | 7.0 [5.9; 7.9] | 5.3 [4.9; 5.8] | <0.001 |
| Postprandial glucose , mmol /l | 9.3 [8.0; 14.1] | 6.1 [5.4; 7.0] | <0.001 |
| Fasting insulin, μIU/ml | 5.3 [3.7; 7.5] | 5.7 [2.2; 9.3] | 0.966 |
| Postprandial insulin , μIU/ml | 15.2 [9.8; 26.1] | 15.4 [8.8; 39.4] | 0.830 |
| HOMA index | 1.6 [1.1; 2.2] | 1.3 [0.6; 2.1] | 0.332 |
| HbA<sub>1c</sub> glycosylated , % | 7.1 [6.5; 8.3] | 6.1 [5.7; 6.5] | <0.001 |
| Total cholesterol, mmol /l | 4.1 [ 3.1; 5.0] | 4.1 [3.2; 5.3] | 0.678 |
| Triacylglycerides , mmol /l | 1.7 [1.2; 2.0] | 1.4 [1.1; 2.2] | 0.332 |
| LDL-C, mmol /l | 2.4 [1.4; 2.9] | 2.4 [1.8; 3.2] | 0.489 |
| HDL-C, mmol /l | 1.0 [ 0.9; 1.3] | 1.1 [ 0.9; 1,2] | 0.801 |
| LDL-C/HDL-C | 1.8 [1.5; 2.9] | 2.2 [1.5; 3.1] | 0.430 |
| TG/HDL-C | 1.6 [1.3; 1.9] | 1.4 [1.0; 2.1] | 0.575 |
| TG/glucose index | 4.0 [ 3.9; 4.1] | 3.8 [3.6; 4.0] | 0.017 |

**Note.** HOMA - Homeostasis Model Assessment; LDL-C: low-density lipoprotein cholesterol; HDL-C: high-density lipoprotein cholesterol; TG - triglycerides; IHD - coronary heart disease; DM - diabetes mellitus.

We found an increase in the absolute content of lymphocytes in the group with a combination of IHD and T2DM (Table 4). The group of patients with IC and T2DM was characterized by a higher absolute content of the total subpopulation of FoxP3+CD4+ lymphocytes and FoxP3+CD25 low lymphocytes (Table 4). After adjusting for sex and BMI, the lymphocyte content was found to be comparable, while the absolute content of FoxP3+CD4+ and FoxP3+CD25 low -lymphocytes and the proportion of FoxP3+CD25 low -cells in patients with DM2 were higher than in patients without DM (Table 4). Higher content of the FoxP3+CD25 low subpopulation, but not of the classic FoxP3+CD25 low Treg lymphocytes in patients with coronary artery disease in the presence of T2DM correspond to changes in autoimmune inflammatory pathologies [8].

**Table 4.** Content of FoxP3+ T lymphocytes in peripheral blood.

| Variables | Patients<br>with a combination of coronary artery<br>disease and T2DM.<br>(n=15] | Patients<br>with coronary artery disease<br>without diabetes.<br>(n=20] | p<br>value |
| --- | --- | --- | --- |
| Lymphocytes, % | 38.2 [34.4; 41.2] | 33.7 [26.3; 45.1] | 0.118 |
| Lymphocytes, ×10<sup>9</sup> /l | 2.6 [2.4; 2.8] | 2.2 [1.7; 2.5] | 0.005 |
| FoxP3+CD4+, % | 3.38 [2.27; 4.23] | 2.28 [1.86; 3.67] | 0.107 |
| FoxP3+CD4+, ×10<sup>7</sup> /l | 4.82 [3.27; 5.73] | 2.93 [1.80; 4.90] | 0.020 |
| FoxP3+CD25<sup>high</sup> , % | 1.69 [1.18; 2.25] | 1.39 [0.95; 2.56] | 0.382 |
| FoxP3+CD25<sup>high</sup> ,<br>×10<sup>7</sup>/l | 2.80 [1.93; 3.28] | 1.87 [1.16; 2.62] | 0.179 |
| FoxP3+CD25<sup>low</sup> , % | 1.66 [1.34; 2.46] | 1.18 [0.81; 1.51] | 0.080 |
| FoxP3+CD25<sup>low</sup> ,<br>×10<sup>7</sup>/l | 1.24 [0.90; 1.77] | 0.84 [0.65; 1.27] | 0.008 |
| Indicators after correction for gender and BMI. |  |  |  |
| Lymphocytes, % | 35.5 [33.0; 36.9] | 37.3 [27.1; 44.4] | 0.598 |
| Lymphocytes, ×10<sup>9</sup> /l | 2.5 [2.2; 2.7] | 2.2 [2.0; 2.5] | 0.133 |
| FoxP3+CD4+, % | 2.72 [1.97; 4.06] | 2.27 [1.76; 2.61] | 0.114 |
| FoxP3+CD4+, ×10<sup>7</sup> /l | 3.53 [2.42; 4.69] | 2.24 [1.80; 3.57] | 0.036 |
| FoxP3+CD25<sup>high</sup> , % | 1.52 [1.05; 2.30] | 1.33 [0.90; 2.38] | 0.419 |
| FoxP3+CD25<sup>high</sup> ,<br>×10<sup>7</sup>/l | 2.21 [1.94; 3.28] | 1.41 [1.09; 2.64] | 0.122 |
| FoxP3+CD25<sup>low</sup>, % | 1.15 [0.98; 1.73] | 0.96 [0.60; 1.15] | 0.046 |
| FoxP3+CD25<sup>low</sup>,<br>×10<sup>7</sup>/l | 1.48 [1.05; 1.97] | 1.07 [0.71; 1.42] | 0.025 |

**Note.** IHD - coronary heart disease; DM - diabetes mellitus; BMI: body mass index; The percentage of cells is indicated from the total number of CD4+ T lymphocytes.

low lymphocytes and in the absolute number of FoxP3+CD25 low lymphocytes was observed. FoxP3+CD25. High lymphocytes with intranuclear localization of FoxP3 were observed compared to patients without diabetes (Table 5). After adjusting for age, the only parameter that distinguished between patients with and without T2DM was the proportion of FoxP3+CD25 low cells with FoxP3 located in the nucleus (Table 5). Thus, for the first time we show not only an increase in the content of these cells, but also a change in their functional potential compared to patients without diabetes.

**Table 5.** Distribution of cells by subcellular localization of FoxP3.

| Indicators | Patients<br>with a combination of coronary<br>artery disease and T2DM.<br>(n=12) | Patients<br>with coronary artery<br>disease without diabetes.<br>(n=22) | P<br>value |
| --- | --- | --- | --- |
| <sup>high</sup> cells with intranuclear localization<br>of FoxP3, % | 98.4 [97.8; 98.7] | 97.8 [95.8; 98.2] | 0.131 |
| FoxP3+CD25 cells<sup>high</sup> with<br>intranuclear localization of FoxP3,<br>×10<sup>7</sup>/l | 7.1 [4.7; 9.7] | 4.2 [2.6; 6.7] | 0.036 |
| <sup>low</sup> cells with intranuclear localization<br>of FoxP3, % | 95.9 [90.0; 97.3] | 85.4 [80.6; 92.0] | 0.004 |
| <sup>low</sup> cells with intranuclear localization<br>of FoxP3, ×10<sup>7</sup>/l | 1.5 [0.6; 2.5] | 0.8 [0.5; 1.0] | 0.048 |
| Indicators after age adjustment. |  |  |  |
| <sup>high</sup> cells with intranuclear localization<br>of FoxP3, % | 99.1 [97.8; 99.6] | 98.0 [96.0; 99.0] | 0.113 |
| <sup>high</sup> cells with intranuclear localization<br>of FoxP3, ×10<sup>7</sup>/l | 5.7 [5.2; 10.6] | 4.2 [3.3; 6.5] | 0.175 |
| <sup>low</sup> cells with intranuclear localization<br>of FoxP3, % | 92.0 [86.4; 95.0] | 88.7 [80.0; 91.4] | 0.040 |
| <sup>low</sup> cells with intranuclear localization<br>of FoxP3, ×10<sup>7</sup>/l | 0.9 [0.7; 1.3] | 0.8 [0.4; 1.1] | 0.152 |

**Note.** IHD - coronary heart disease; DM - diabetes mellitus; The percentage of cells is indicated from the total number of FoxP3+CD25<sup>high</sup> - and FoxP3+CD25<sup>low</sup> - lymphocytes, respectively.

Patients with a combination of coronary artery disease and T2DM were characterized by an increase in the serum content of the chemokine CCL22 (CC Motif Chemokine Ligand 22) and TNF-α, as well as an increase in the content of CCL22 in the supernatant of the daily intake. LPS-stimulated mononuclear leukocyte cultures, indicating the contribution of peripheral blood cells to changing the systemic concentration of this chemokine.

In the general group of patients with coronary heart disease we found a moderate relationship between the concentration of CCL22 in the supernatant of daily cultures of mononuclear leukocytes stimulated with LPS and the values of the TG/glucose index (Rs = 0.620; p = 0.024), as well as between the relative amount of high -FoxP3 + CD25 lymphocytes in peripheral blood and the serum concentration of CCL22 (Rs =0.587; p=0.044).

Only in patients without diabetes was there a trend for the Gensini score to correlate with the relative number of FoxP3+CD25 high cells, Treg with the intranuclear location of FoxP3 (Rs =-0.471; p=0.076). When coronary artery disease is combined with T2DM, the strongest relationship was found between the Gensini Score and the absolute number of lymphocytes (Rs =0.564; p=0.015), as well as HDL-C content (Rs =-0.515 ; p=0.029)..

We divided the entire group of patients with coronary heart disease into subgroups based on the Gensini Score tertiles : first tertile (Gensini Score <17 points; low Gensini Score values); second tertile (Gensini score 17 to 45 points; average values of the Gensini score); third tertile (Gensini Score >45 points; high Gensini Score values). It should be noted that the tertiles of our sample were similar to those of large-scale studies [11].

In patients with DM2 and without DM, the values of the TG/glucose index differed (in patients in the first tertile) (Table 6), while there were no intergroup differences in the metabolic parameters depending on the value of the Gensini Score in patients with DM (Table 6).

**Table 6.**
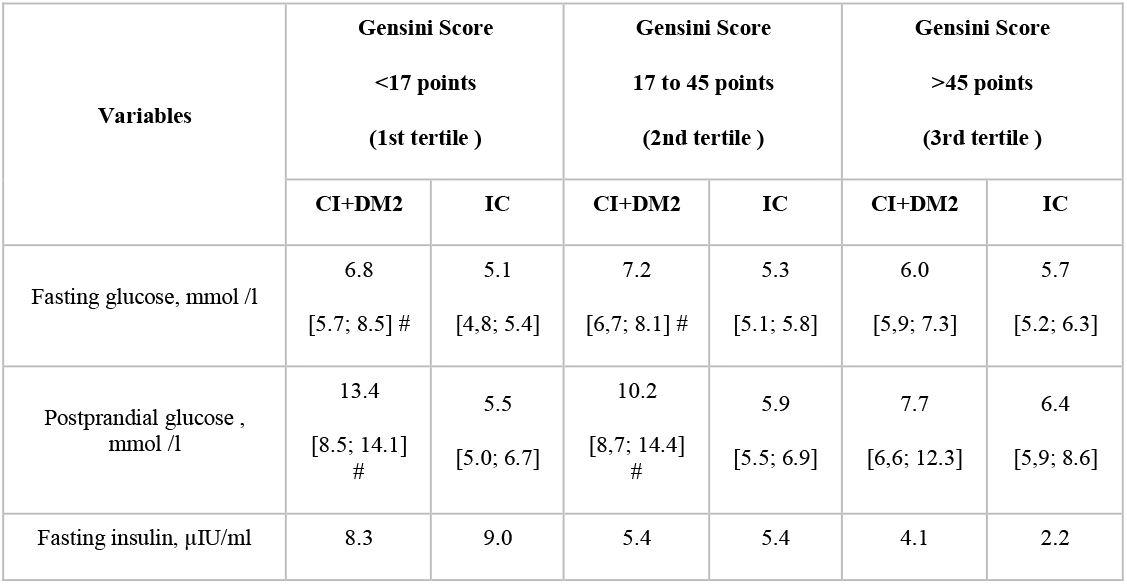
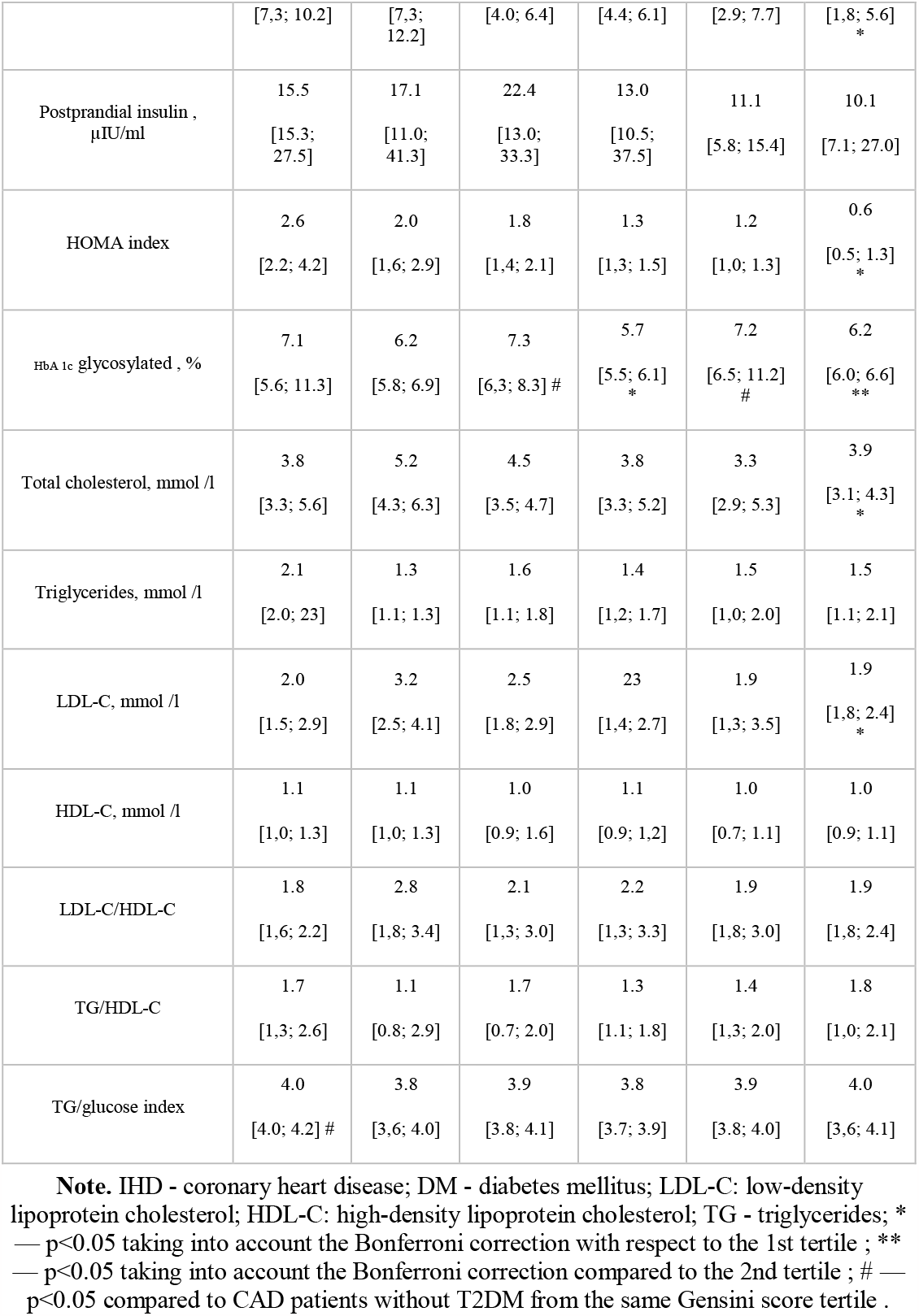
Parameters of carbohydrate and lipid metabolism according to the Gensisni Score value in patients with and without type 2 diabetes mellitus.

When analyzing data from flow cytometry and flow cytometry with visualization, it turned out that patients with coronary artery disease and T2DM with average values of Gensini Score (second tertile of Gensini Score) were characterized by a tendency to increase the absolute content by FoxP3+. CD25 high and FoxP3 + CD25 low T cells with FoxP3 located in the nucleus, compared to patients with low Gensini Score values, as well as an increase in the absolute content of FoxP3+CD25 low T lymphocytes, the content of FoxP3+CD25 high and FoxP3+CD25 low T cells with FoxP3 transfer to the nucleus and relative content of FoxP3+CD25 low, T cells with nuclear transfer of FoxP3 compared to patients without diabetes (Table 7). In patients with average Gensini Score values without diabetes, we found a trend towards a decrease in the relative content of FoxP3+CD25 low T lymphocytes compared to patients with low Gensini Score values (Table 7).

**Table 7.** Characteristics of FoxP3+CD25 high - and FoxP3+CD25 low - cells according to the Gensisni Score value in patients with and without type 2 diabetes mellitus.

| Parameter | Gensini score <17 points<br>(1st tertile ) |  | Gensini obtains between 17 and 45 points<br>(2nd tertile ) |  | Gensini Score >45 points<br>(3rd tertile ) |  |
| --- | --- | --- | --- | --- | --- | --- |
|  | CI+DM2 | IC | CI+DM2 | IC | CI+DM2 | IC |
| FoxP3+CD4+, % | 2.6<br>[2,4; 2.7] | 23<br>[2.0; 2.7] | 2.4<br>[1.6; 4.6] | 2.1<br>[1,7; 2.6] | 3.6<br>[2.8; 4.9] | 1.7<br>[0.6; 2.7] |
| FoxP3+CD4+, ×10<sup>7</sup>/l | 2.4<br>[2.2; 3.7] | 2.1<br>[1.8; 2.8] | 3.5<br>[2.5; 7.3] | 2.9<br>[2.0; 3.7] | 4.1<br>[3.4; 5.2] | 2.4<br>[1.0; 4.5] |
| FoxP3+CD25<sup>high</sup>, % | 1.5<br>[1.1; 1.9] | 1.3<br>[1.0; 1.6] | 1.5<br>[1.0; 2.1] | 2.1<br>[1.4; 3.1] | 2.0<br>[1.5; 2.7] | 1.2<br>[0.5; 1.8] |
| FoxP3+CD25<sup>high</sup>, ×10<sup>7</sup>/l | 1.9<br>[1.3; 3.1] | 1.3<br>[1.0; 1.6] | 2.9<br>[2.0; 4.0] | 2.0<br>[1.2; 3.9] | 2.1<br>[2.0; 3.0] | 1.7<br>[1.0; 3.1] |
| FoxP3+CD25<sup>low</sup>, % | 1.1<br>[1.1; 1.2] | 1.3<br>[1.0; 1.2] | 1.4<br>[1.0; 1.7] | 0.6<br>[0.5; 1.1] * | 1.4<br>[1.0; 1.9] | 0.6<br>[0.4; 0.8] |
| FoxP3+CD25<sup>low</sup>, ×10<sup>7</sup>/l | 1.1<br>[1.0; 1.1] | 1.3<br>[0.9; 1.4] | 1.7<br>[1.4; 2.1] # | 0.8<br>[0.6; 0.9] | 1.7<br>[1.4; 2.4] | 1.0<br>[0.6; 1.6] |
| <sup>high</sup> cells with intranuclear localization of FoxP3, % | 98.7<br>[97.8; 99.9] | 98.9<br>[98.4; 99.2] | 99.6<br>[99.1; 99.6] | 97.9<br>[95.8; 99.7] | 98.6<br>[97.9; 99.3] | 97.9<br>[97.3; 98.2] |
| <sup>high</sup> cells with intranuclear localization of FoxP3, ×10<sup>7</sup>/l | 5.0<br>[1.4; 5.3] | 3.7<br>[2.8; 5.9] | 7.6<br>[7.0; 13.5] *# | 6.2<br>[2.2; 6.8] | 5.3<br>[2.7; 14.1] | 3.7<br>[3.4; 5.4] |
| <sup>low</sup> cells with intranuclear localization of FoxP3, % | 82.8<br>[79.7; 91.5] | 88.1<br>[84.3; 90.0] | 94.7<br>[92.7; 95.3] # | 88.7<br>[70.2; 91.4] | 92.4<br>[74.5; 100] | 90.2<br>[85.1; 92.1] |
| <sup>low</sup> cells with intranuclear localization of FoxP3, ×10<sup>7</sup>/l | 0.7<br>[0.6; 0.7] | 1.1<br>[0.8; 1.2] | 1.3<br>[1.2; 1.3] *# | 0.8<br>[0.3; 0.9] | 0.8<br>[0.6; 2.9] | 0.5<br>[0.4; 1.1] |

Note. IHD - coronary heart disease; DM - diabetes mellitus; \* — p<0.1 taking into account the Bonferroni correction compared to the 1st tertile ; #- p<0.05 compared to patients with CAD without T2DM from the same Gensini score tertile .

## DISCUSSION

Most of the information on the functioning of FoxP3+ regulatory T cells is based on an assessment of their percentage and does not take into account the subpopulation of FoxP3+CD25 cells [5]. In our pilot study, we identified for the first time an increase in the absolute content of FoxP3+ T cells in patients with a combination of coronary heart disease and T2DM compared to patients with IHD without carbohydrate metabolism disorders. This was primarily due to an increase in the content of the FoxP3+ CD25 lymphocyte subpopulation, which has received little attention to date.

Ferreira RC et al. (2017) demonstrated that these cells share similar characteristics with Treg cells, such as demethylation of FoxP3, constitutive expression of the transcription factor Helios (a marker of Treg of thymic origin) in most cells, and the inability to produce the cytokine IL-2. 8]. The difference with conventional CD127 or CD25 high FoxP3+ Treg cells was the reduced expression of Treg- associated molecules (FoxP3 and CTLA-4). From which the authors concluded that these cells are the final stage of the life cycle of Treg lymphocytes. Furthermore, chronic stimulation during active autoimmune inflammation leads to an increase in their number [8].

In accordance with our data, a more pronounced low-intensity chronic inflammation in T2DM, which is manifested in the examined patients with coronary artery disease by an increase in the serum concentration of TNF-α, is also associated with an increase in the absolute number of FoxP3+. CD25 low -lymphocytes in the peripheral blood and is accompanied by an increase in the transfer of FoxP3 to their nucleus. Previously, there was no information about an increase in the content of this cellular subpopulation in patients with T2DM. At the same time, it has been shown that patients with T2DM are characterized by a decrease in the relative content of CD4+CD25+FOXP3+ Treg cells [12]. Another group of researchers found that Treg lymphocyte content decreases with a duration of T2DM of more than 10 years [13]. The median duration of diabetes in the patients included in our study was less than 10 years, but exceeded 5 years. It can be assumed that the increase in the number of FoxP3+CD25 low lymphocytes that we detected is an intermediate stage and indicates the exhaustion of the regulatory potential of the immune system. We believe that an increase in the content of FoxP3+CD25 low lymphocytes can be considered as a promising cellular marker of low-intensity chronic inflammation in patients with a combination of IC and T2DM.

We have demonstrated for the first time in Latin America that the identified changes are more typical of patients with moderately severe atherosclerosis. At the initial stages of the development of atherosclerosis, with low Gensini values, the content of FoxP3+ lymphocytes and the level of intranuclear content of FoxP3 in them were comparable to those of patients without diabetes. It is likely that at this stage of disease development the mobilization and activation of T lymphocytes is not yet sufficiently manifested. We did not detect significant differences with high Gensini Score values, probably because the mobilization reserve and the ability to activate the T-regulatory link begin to be depleted. To confirm this hypothesis, it is necessary to study the cellular composition of atherosclerotic plaques, which was not possible in this group of patients with stable coronary artery disease. It is noteworthy that lymphocytes increase as the severity of atherosclerosis increases, indicating fundamental differences in the state of the regulatory mechanisms of the immune response in the presence of disorders of carbohydrate metabolism in patients with coronary artery disease.

One of the probable pathogenic links that contribute to the mobilization of FoxP3+ lymphocytes in patients with DM2 may be a change in the production of the chemokine CCL22, the concentration of which increased in the serum and in the supernatant of leukocyte cultures stimulated with LPS in the presence from DM. CCL22 is known to be the main chemoattractant of FoxP3+ Treg cells [14▪]. The level of insulin resistance may have a direct impact on CCL22 production, given the relationship we found between LPS-stimulated CCL22 production by mononuclear leukocytes and the TG/glucose ratio in patients. A low frequency of Treg cells was shown to be associated with the destabilization of atherosclerotic plaques in experimental animals [15]. It can be assumed that in T2DM there is a CCL22- mediated alteration in the recruitment of Treg lymphocytes to the atheroma, which ultimately contributes to Treg deficiency in the vascular wall, increased inflammation and the progression of atherosclerosis. [6]. This assumption is also at the level of a hypothesis and requires confirmation in future studies.

### Limitations of the study

Limitations of the study include the small patient sample size and its observational, cross-sectional design. This pilot study confirmed the need to study the subpopulations of high FoxP3+CD25 lymphocytes and low FoxP3+CD25 lymphocytes in a larger group of patients. Furthermore, a future prospective study would allow us to conclude that FoxP3+CD25 low lymphocytes can be identified as a biomarker associated with a favorable or unfavorable prognosis in patients.

The results obtained on the different content of FoxP3+CD25 low lymphocytes in patients with DM2 depending on the Gensini score do not exclude the possibility that treatment tactics for patients with DM2 at different stages of the development of atherosclerosis may differ. It is known that several hypoglycemic drugs can affect the functioning of the immune system. The vast majority of patients with T2DM in our study received hypoglycemic treatment, including metformin (86.4% of patients). The available data on the presence of immunomodulatory effects of metformin are contradictory. Several studies have found its stimulatory effect on FoxP3 expression [16], while in cancer patients, administration of metformin, on the contrary, caused a decrease in the content of FoxP3+ T lymphocytes [17▪]. It is advisable to conduct additional studies aimed at studying the effect of biguanides on the properties of FoxP3+ T cells in patients with a combination of coronary artery disease and T2DM.

## CONCLUSION

Patients with a combination of CHF and T2DM are characterized by an increase in the relative and absolute content of FoxP3+CD25 lymphocytes in the peripheral blood and an increase in the intranuclear content of FoxP3 in them, which is more pronounced in patients with disease moderate. atherosclerosis (Gensini score 17 to 45 points). The identified changes are associated with an increase in the production of the chemokine CCL22 and the degree of insulin resistance and do not exclude in the future the possibility of developing different approaches for the treatment of patients with coronary artery disease and T2DM, depending on the severity of coronary atherosclerosis and the content of FoxP3 +CD25 low cells.

## Data Availability

All data produced in the present work are conteined in the manuscript

